# Leftover Infant Milk After Bottle Feeding: Parental Practices and Microbiological Findings

**DOI:** 10.64898/2026.02.13.26346179

**Authors:** Anna Zychlinsky Scharff, Ludwig Sedlacek, Ariana de Oliveira Mekonnen, Ioannis Liolios, Simon Ritter, Franziska Fuchs, Christine Happle

**Author notes:** Corresponding Author: Anna Zychlinsky Scharff, MD, Molecular Pharmacology Program, Memorial Sloan Kettering Cancer Center, 1275 York Avenue, ZRC-18 South, New York, NY 10065, USA.

## Abstract

**Importance:** Current guidelines from the World Health Organization, Centers for Disease Control and Prevention, and Academy of Breastfeeding Medicine recommend discarding all milk remaining in bottles immediately after infant feeding. However, these recommendations lack supporting microbiological evidence from studies of actual infant feeding, imposing substantial financial and emotional burden on the 78 million families worldwide who bottle-feed their infants.

**Objective:** To determine (1) the financial, emotional, and time burden associated with bottle feeding and parental milk disposal practices, and (2) bacterial growth in leftover human milk and formula under different storage conditions.

**Design:** (1) Cross-sectional online survey (January 2023-February 2024) and (2) prospective microbiological cohort study.

**Setting:** (1) Online survey, (2) infants recruited in Hannover, Germany

**Participants:** (1) Survey respondents (n=1056; 99% mothers) and (2) healthy, full-term, bottle-fed infants (n=44; 17 humanmilk, 27 formula) aged 0-12 months.

**Main Outcomes and Measures:** Parental burden scores, milk disposal frequency, and bacterial colony-forming units (CFU)/ml in milk samples before feeding, immediately after feeding, and at 4, 8, and 24 hours post-feeding at 4°C and 20°C.

**Results:** Among surveyed parents, 46% discarded leftover milk daily, yet 84% reported they would keep milk longer if deemed safe. In microbiological testing, median bacterial burden in humanmilk increased from 4200 CFU/ml (range 300-350,000) pre-feeding to 24,600 CFU/ml (range 1900-29,004,400) post-feeding, but showed no significant further increase at 4 hours (p=0.82) or 8 hours (p=0.64) when stored at either 4°C or 20°C. Formula showed similar stability: median CFU/ml increased from 0 (range 0-10,700) to 11,700 (range 1900-630,000) post-feeding, with no significant change at 4 hours (p=0.91) or 8 hours (p=0.73) at either temperature. Significant bacterial growth occurred only after 24 hours at 20°C (p<0.001).

**Conclusions and Relevance:** Bacterial burden in leftover infant milk remained stable below concerning thresholds for 8 hours when refrigerated and 4-8 hours at room temperature, challenging current guidelines that mandate immediate disposal. Evidence-based guideline revision could reduce financial burden and milk waste for families around the globe without compromising infant safety.

**Key Points:** *Question:* How long is it safe to offer leftover milk in a bottle to an infant that has previously drunk from it?

*Findings:* The number of bacteria in leftover human milk or formula did not significantly increase from 0 to 8h post-feeding in milk bottles sampled from 44 infants, regardless of whether the milk was kept at room temperature or refrigerated.

*Meaning:* Leftover milk may be safely reoffered beyond the limits of the current guidelines.

## Introduction

Globally, more than half of infants receive some bottle feeding before six months of age^1^. While many studies focus on increasing breastfeeding rates through targeted interventions, data on supporting bottle-feeding families is scarce^2–5^.

The World Health organization (WHO), the Centers for Disease Control and Prevention (CDC), the Academy of Breastfeeding Medicine and European authorities issue strict and specific guidelines regarding the preparation and storage of infant milk. These recommend that, once an infant has been fed from a bottle of humanmilk, residual milk should be discarded after 2 hours^6^. For formula, the standard is even stricter, requiring leftover milk to be discarded immediately^7^. However, little research has explored the stringency with which parents and caregivers implement such protocols, and real-world data supporting or contradicting these recommendations are scarce^8–11^. The Academy of Breastfeeding Medicine notes that “The length of time the milk can be kept at room temperature once the infant has partially fed from the cup or bottle would theoretically depend on the initial bacterial load in the milk, how long the milk has been thawed, and the ambient temperature. There has been insufficient research done to provide recommendations in this regard. However, based on related evidence thus far, it seems reasonable to discard the remaining milk within 1–2 hours after the infant is finished feeding”^12^. While a conservative, risk-averse approach makes sense in the context of infant safety, the drawbacks are under-studied. The effort expended to pump human human milk is significant, and likewise formula costs time and money to obtain^13,14^. The formula shortage in the United States in 2022 highlighted the need for more data in this area, as parents were forced to make difficult decisions regarding infant feeding safety protocols^15^.

To address the burdens associated with pumping human milk and purchasing formula, as well as milk storage habits, we conducted a survey of 1056 parents. Furthermore, to generate evidence on bacteria levels in bottled milk after feeding, we collected samples from the mouths and milk bottles of 44 infants (27 formula-fed and 17 human milk-fed) and assessed bacterial growth at different temperatures over time.

## Materials and Methods

### Survey

To explore the experiences of parents and caregivers regarding bottle feeding, we conducted a cross-sectional study using a convenience sample collected via an online questionnaire. Inclusion criteria were (1) being a parent or guardian to a child 36 months or younger and (2) consenting to participate in an online survey. The study was conducted from June to October 2023. Survey participants were recruited with internet links and flyers. Data were collected anonymously, and survey participants were able to skip questions they preferred not to answer. Only parents that reported bottle-feeding were asked to specify information on their bottle-feeding practices. Hence, the total number of responses per question varied. For some items, e.g. financial or emotional impact of infant feeding in their setting, participants were asked for their assessment on a Likert scale from 1-10^16^. Parents with multiple children were asked to report on their practices regarding their youngest infant, so that all respondents were reporting on practices occurring within the previous 36 months. While nomenclature differs, we https://research.sahmri.org.au/en/persons/maria-makrides defined breastfeeding/chestfeeding as infant feeding directly from the lactating parent’s nipple^17^. Pumped human milk refers to milk extracted from the lactating parent by any method, including manual or electric pumps and/or hand expression.

### Infant recruitment and sampling

Healthy bottle-fed infants between the age of 0 and 12 months whose parent or guardian consented to participation in this study were brought to the study center for a feeding session. No antibiotic treatment of infant or lactating parent in the previous 30 days was a requirement for inclusion in the study. Caregivers received prior instructions to ensure best practices in bottle preparation and were asked to mix their formula according to manufacturer’s instructions. For pumped human milk, we asked caregivers to provide milk pumped within the previous 24h and kept in the refrigerator until feeding, or frozen human milk defrosted within 2 hours prior to feeding. To ensure feeding comfort for the infant, families brought the bottle of their choice (Suppl Fig 1a). During the study visit, we first collected a buccal swab to identify the oral flora of the infant. Next, using sterile technique, we collected a pre-feeding milk sample from the prepared bottle. The caregiver was then invited to feed the infant at leisure, and a post-feeding sample of the milk left in the bottle after infant satiety was collected, again under sterile conditions. The length of feeding time was recorded. We contacted parents 7 days after sample collection to enquire about any subsequent illness and/or symptoms. All infants remained healthy in the week following their study visit.

### Microbiological Testing

Samples were immediately transported to the on-site microbiology laboratory. Oral swabs were cultured on Columbia blood agar (aerobic conditions) and chocolate blood agar (5% CO_2_). Plates were incubated for 48 h and read at 24 and 48 h. From each milk sample, a pure aliquot and serial dilutions were plated on Columbia blood agar immediately upon arrival in the lab (timepoint 0). Samples and their dilutions were then split and incubated at both 4°C and 20°C for 4, 8 and 24 hours respectively. Colony forming units (CFU) and species were determined for each time point by plating serial dilutions on Columbia blood agar. Bacterial species were identified by Matrix Assisted Laser Desorption/Ionization Time of Flight (MALDI TOF) Mass Spectrometry analyses, as standardized for medical microbiology laboratories (standard DIN EN ISO 15189). The time points of 4 and 8 hours of incubation were chosen to reflect likely real-world use of leftover milk, and 24 h was employed as a timepoint when bacterial growth at 20°C was expected to increase substantially. The limit of detection of our assay was 100 CFU/ml.

### Data Analysis

Statistical analyses were carried out using R (Version 4.4.0). Burden scores from the online survey were treated as continuous variables. As these variables violated normality (Shapiro-Wilks test, p < 0.001 for all distributions) a non-parametric approach was used. For comparisons between two independent groups, the Wilcoxon rank-sum test was applied. For multiple independent groups, the Kruskal–Wallis test was first performed to assess overall differences in distributions. Pairwise Wilcoxon rank-sum tests were then conducted within the same category, with p-values adjusted using the Holm-Bonferroni correction for multiple comparisons. The effect of categorical variables was assessed via Chi Square testing.

We analyzed differences in bacterial growth between group medians using the Mood’s Median Test. Mood’s Median Test was chosen to compare central tendency in CFU distributions due to extreme skewness and outliers in bacterial count data. We applied the Holm-Bonferroni correction in the case of multiple testing.

### Research Ethics

The study protocol was approved by the research ethics board (Hannover Medical School # Nr. 10492_BO_K_2022). Parents of all infants donating milk samples gave their written informed consent.

## Results

In total, n=1056 parents responded to the online questionnaire (99% identified as mothers, 1% as fathers). Further characteristics of the population are summarized in Table 1a. Sixty-one percent (645/1053) of parents reported partial or exclusive bottle feeding (Suppl. Fig 2). We asked respondents to rate the challenges related to nursing, bottle-feeding pumped human milk, and bottle-feeding formula on a Likert scale of 0 (very easy) to 10 (very difficult). Bottle feeding of pumped humanmilk was consistently associated with higher difficulty ratings than nursing across all four burden domains (all p<0.001). Formula feeding showed higher financial and logistical burden than nursing (all p<0.001), but was associated with lower emotional and time burden (p<0.001).

**Table 1a:** Cohort characteristics of survey respondents.

| **Sex of Parent** |  |  |
| --- | --- | --- |
| *Information available in n=1046* | n= | % |
| Male | 15 | 1 |
| Female | 1031 | 99 |
| **Marital status** |  |  |
| *Information available in n=1050* | n= | % |
| Partnered | 1000 | 95 |
| Single | 50 | 5 |
| **Number of children** |  |  |
| *Information available in n=1056* | n= | % |
| 1 | 603 | 57 |
| 2 | 335 | 32 |
| 3 | 86 | 8 |
| 4 | 23 | 2 |
| 5 | 4 | <1 |
| 6+ | 5 | <1 |
| **Age of youngest child at breastfeeding cessation** |  |  |
| *Information available in n=886* | n= | % |
| <3 months | 124 | 14 |
| 3-6 months | 86 | 10 |
| 6-12 months | 90 | 10 |
| Over 12 months | 108 | 13 |
| Currently breastfeeding | 478 | 53 |
| **Migration background** |  |  |
| *Information available in n=1458* | n= | % |
| Yes | 51 | 5 |
| No | 999 | 95 |
| **Mode of milk feeding** |  |  |
| *Information available in n=1053* | n= | % |
| Exclusively nursing | 408 | 39 |
| Exclusively bottle feeding | 156 | 15 |
| Combination feeding | 489 | 46 |
| **Sex of child** |  |  |
| *Information available in n=1055* | n= | % |
| Male | 505 | 48 |
| Female | 550 | 52 |
| **Age** |  |  |
| *Information available in n=1050* | Median | IQR |
| Parent's current age (years) | 34 | 6.75 |
| Parent's age at first birth (years) | 31 | 6 |
| Age of youngest child (months) | 12 | 16 |

**Table 1b:** Cohort characteristics of infants in microbiological milk testing cohort.

| **Sex of infant** | **n=** | **%** |
| --- | --- | --- |
| Male | 21 | 48 |
| Female | 23 | 52 |
| **Type of milk** | **n=** | **%** |
| Formula | 27 | 61 |
| Human milk | 17 | 39 |
| **Age, drinking characteristics** | *Median* | *IQR* |
| Age of infants (months) | 6.21 | 5.31 |
| Time drinking from bottle (min) | 7 | 4 |
| Milk consumed (ml) | 110 | 50 |

Of bottle-feeding parents, 46% (268/585) reported discarding leftover milk at least once per day (Figure 2a). However, most parents (84%, 499/591) would prefer to discard less milk if they could confidently do so without compromising the health and safety of their child (Figure 2B).

**Figure 1.**
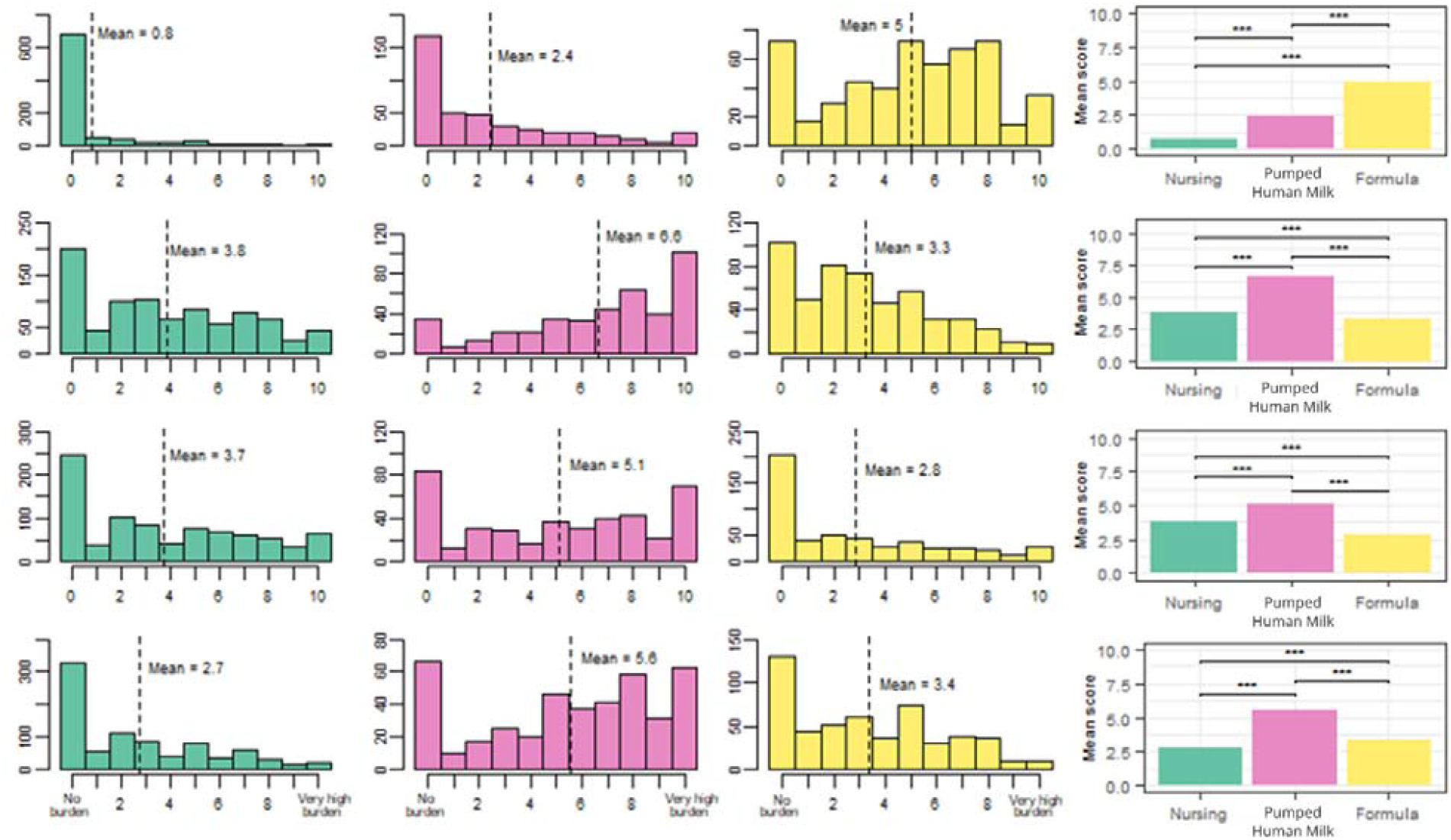
Parental Financial, Time, Emotional and Logistical Burdens by Infant Feeding Method Figure 1: Frequency distributions (first three columns) of surveyed parents reporting difficulty scores (x-axis) for nursing (green), bottle-feeding pumped human milk (pink) and bottle-feeding formula (yellow). Parents were queried for financial (top row), time (second row), emotional (third row) and logistical burden (bottom row) of each feeding modality. Composite graphs (last column) show mean values and significant differences (***p<0.001). Tests used: A Shapiro-Wilks test was employed to assess the non-normality of the data (p<0.001). A Kruskal-Wallis test was then used to assess overall difference in distributions (p < 0.001). Subsequently, pairwise Wilcoxon rank-sum tests were performed within the same burden category, with p-values adjusted using the Holm-Bonferroni correction.

**Figure 2.**
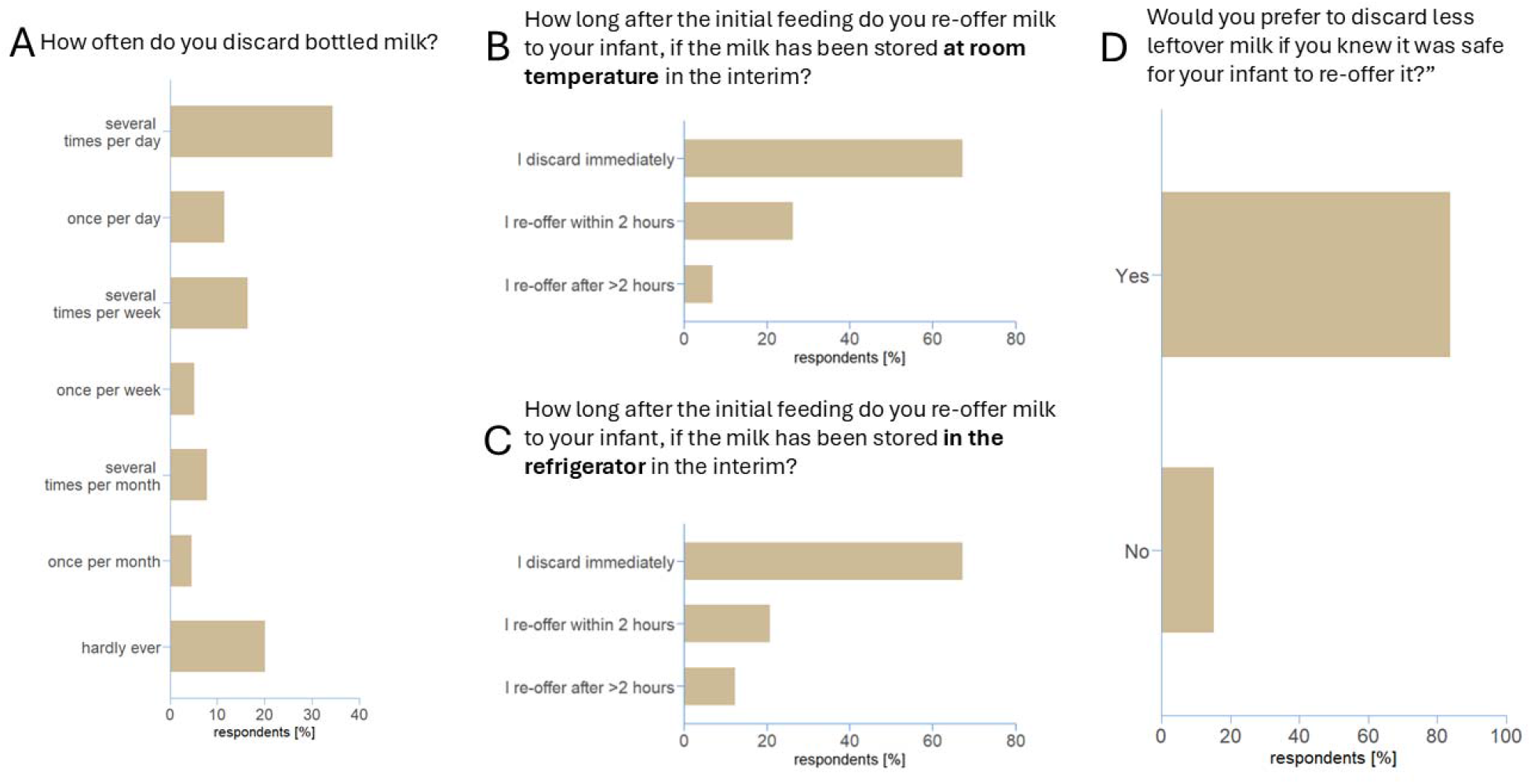
Parental Management and Disposal of Leftover Milk After Infant Feeding Figure 2: Parents practices and preferences regarding re-offering milk leftover after a feeding. (A) Frequency of leftover milk disposal. Parental practices regarding leftover milk if kept at room temperature (B), or in refrigerator (C). Percentage of parents indicating a preference for reducing disposal of leftover milk (D)

Next, we explored the choices of parents regarding timing of leftover milk disposal. Sixty-seven percent of those surveyed choose to discard leftover milk immediately (428/638, Fig 2c, 2d). Only a small minority, 7% (43/638) reoffered leftover milk after more than 2 hours of storage at room temperature (Fig 2c). Most of these parents (75%, 30/40) offered the leftover bottle to their infants within six hours (Suppl. Fig 3a). When milk was refrigerated, a larger, but still small cohort of parents [12% (78/638)] felt comfortable reusing the milk (Fig 2d).

Based on this survey data, we reasoned that discarding milk presents a relevant problem for families. We set out to correct the dearth of evidence on bacterial levels in human milk and formula after first use, to aid in decision making regarding the disposal of leftover milk.

To this end, we recruited 44 bottle-fed infants for microbiological analysis of the milk leftover after a feed. We included 21 infants with formula bottles and 17 infants with pumped human milk in their bottles in our primary analysis. Six infants in the formula group were excluded for protocol deviations. In three cases, the parents indicated on the questionnaire accompanying the study visit that they had not complied with the study requirement to follow the manufacturer’s preparation instructions. In Germany, the instructions recommend boiling water, then cooling it partially and preparing the formula, a practice followed by most surveyed parents (Supp Fig 4a). A further three infants were given formula containing probiotic cultures, confounding the microbiological tests. The median duration of a feeding session in our study was 7.48 min (Suppl Fig 1b). We found that longer feeding was not significantly associated with higher bacterial CFU in the post-feeding milk (Suppl Fig 5a and 5b). The bottle material did not affect bacterial counts (Suppl. Fig 6), nor did infant sex or age (Suppl. Fig 7a and 7b).

For human milk, prior to the feeding, the median CFU/ml was 4200 (Range 300-350,000). Immediately after the infant finished, the CFU/ml increased to a median of 24,600 (Range 1900-29,004,400) (Figure 3A). After 4h of incubation at 4°C or 20°C, the median bacterial burden in both pre- and post-feeding human ilk remained stable. After 8h storage at either temperature, bacterial counts in pre-feeding human milk sample cultures remained within baseline ranges. Post-feeding human milk samples at both temperatures increased beyond the range of timepoint 0, but this increase was not statistically significant. At the late time point (24h), we observed robust, albeit not significant bacterial growth at 20°C (Fig 3A).

**Figure 3.**
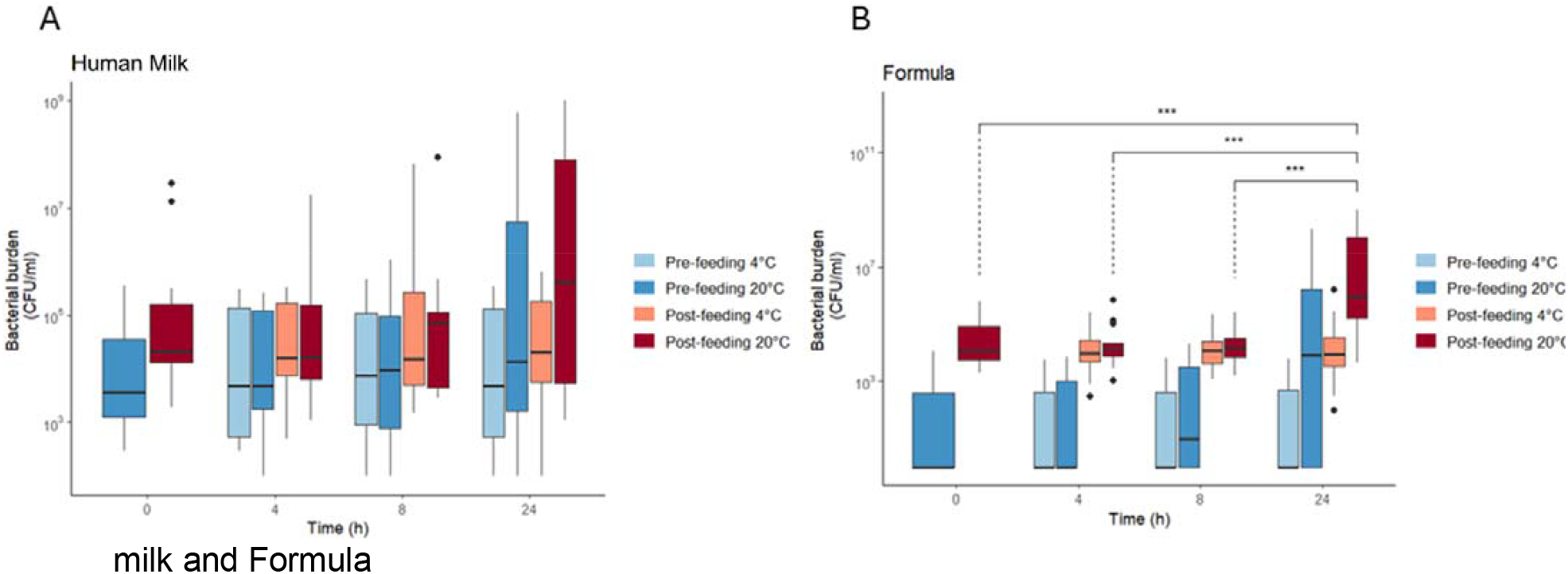
Impact of Storage Time and Temperature on Bacterial Growth in Human milk and Formula Figure 3: Bacteria levels in human human milk and formula under different storage conditions. (A) CFU/ml in human milk prior to (blue) and after (red) bottle feeding and following 4, 8 and 24h incubation at 4°C (lighter shades) or 20°C (darker shades). (B) CFU/ml in formula prior to (blue) and after (red) bottle feeding and following 4, 8 and 24h incubation at 4°C (lighter shades) or 20°C (darker shades). Bars display median plus interquartile range, CFU: colony forming units, h: hours, °C: degrees Celsius, *** p<0.001). No significant differences were found in (A). Test used: Pairwise Mood’s median tests were conducted across all subgroups, with p-values adjusted using the Holm-Bonferroni correction.

Formula, predictably, had lower baseline pre-feeding bacterial load than human milk. Pre-feeding, the median was 0 CFU/ml (Range 0-10,700), rising to a median of 11,700 CFU/ml (range 1900-630,000) after feeding (Fig 3b). There were no significant differences between timepoints 0, 4, and 8h, neither after storage at 4°C nor at 20°C, for pre- and post-feeding formula. A significant increase in bacterial burden occurred only for post-feeding formula after 24h storage at 20°C (p<0.001; Fig 3B).

Next, we investigated the source of bacteria in post-feeding milk. We analyzed buccal swabs of each infant and compared these to the bacterial composition in their respective milk samples. Interestingly, we found that in human milk samples, most bacterial species identified post-feeding had already been present in the pre-feeding sample (Fig. 4a, Suppl. Figure 8). In contrast, in formula, most post-feeding bacterial species mirrored the infants’ own oral flora (Figure 4b, Suppl. Figure 8). Most bacterial species in milk samples could be traced back to either the milk sample before feeding or the oral cavity of the infants. Notably, the source of bacterial species in post-feeding milk varied widely between individuals (Figure 4, Suppl. Figure 9a,9b).

**Figure 4.**
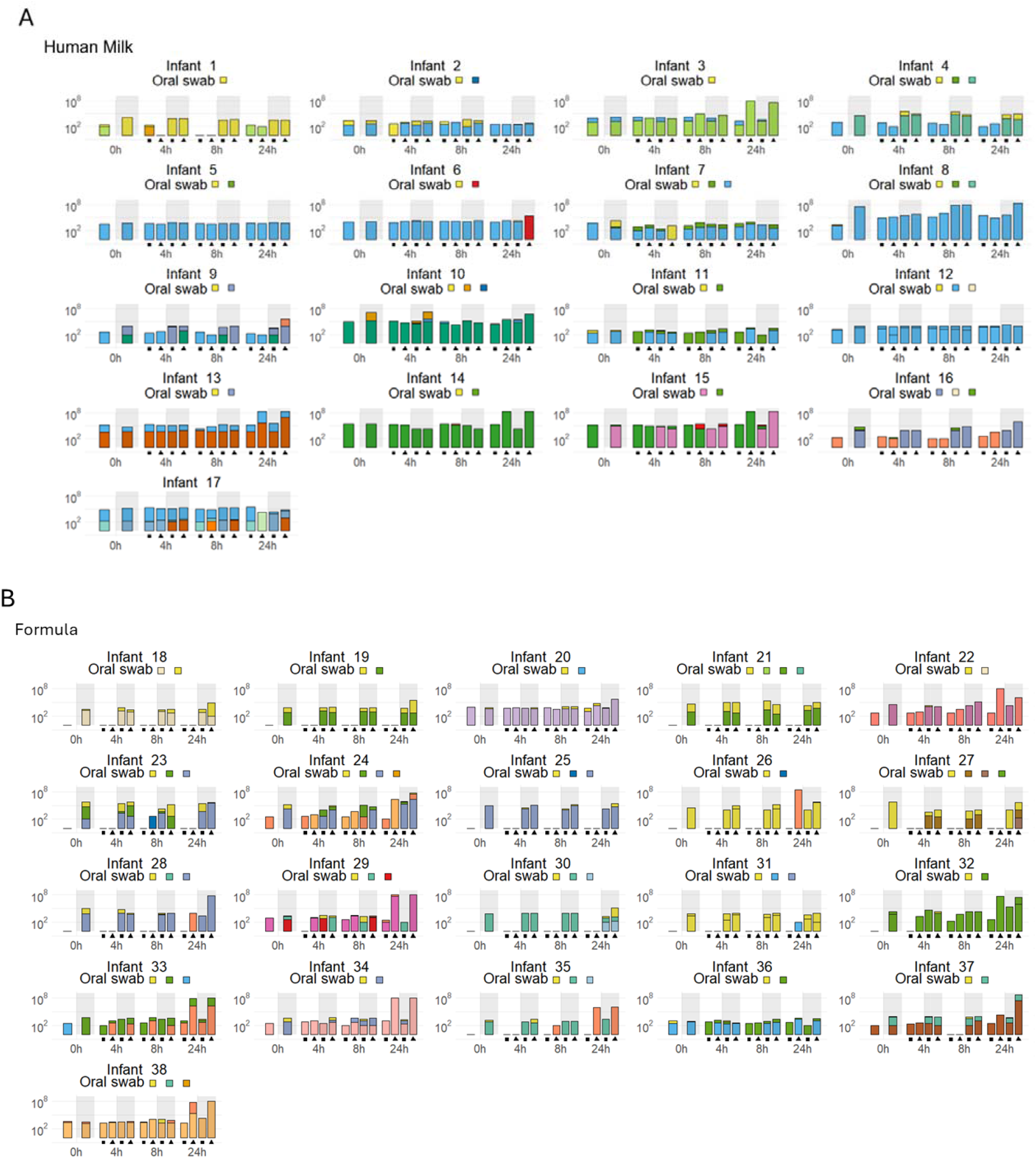
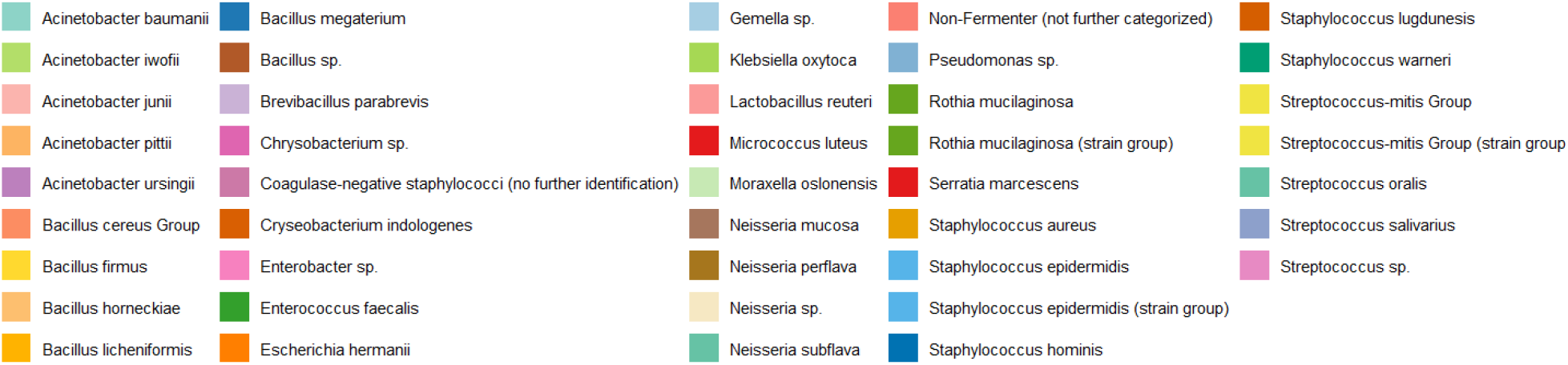
Bacterial Profiles of Human Milk and Formula Over Time After Feeding Bacterial species composition of milk samples over time after infant feeding. (A) Bacterial species composition of human milk samples from individual infants (n=17) at 0 h (directly after feeding), 4 h, 8 h, and 24 h post-feeding. Samples collected from bottle prior to feeding are shown in the grey zone; samples collected from bottle after feeding are shown in the white zone. Incubation at 4 °C is indicated by square symbol (■), incubation at 20 °C by triangle symbol (▲). Column colors denote bacterial species (see color key). Where multiple bacterial species were identified, their relative abundance is reflected by the proportional area of each color within a bar. Bacterial species detected in infant oral swabs prior to feeding are displayed qualitatively, with all identified species shown in color boxes under the “oral swab” label. (B) Equivalent analysis for formula-fed infants (n=21).

In human milk samples, the most common bacterial species by far were *Staphylococcus epidermidis* and *Streptococcus mitis*, with 88% and 65%, respectively, (Figure 4a, Suppl. Table 1), which is broadly consistent with previous reports on the microbiome of human milk^18–24^. *Streptococcus mitis* was also present in 92% infant buccal swabs. In all infants consuming formula bottles, at least one sample was *Streptococcus mitis* positive, illustrating the widespread presence of this commensal of the oral flora in human milk bottles (Figure 4a, Suppl Table 2). Similarly, existing literature has established the presence of *Staphylococcus aureus* in a relevant subset of human milk samples, and we recapitulate this finding (Figure 4a, Suppl. Table 2)^25–31^.

Our survey data indicated that 12% of parents prepared formula with tap water that had not been boiled (Suppl Figure 4a). We therefore examined the bacterial load of the formula inadvertently made with unboiled water in 3 infants (Suppl. Figure 10a). Lack of boiling increased bacterial levels in pre-feeding milk, but the post-feeding milk bacterial burden remained remarkably similar to that of post-feeding formula made with boiled water.

Despite our instructions, three participants used probiotic-containing formula, and were thus excluded from the primary analysis. Predictably, we found large quantities of the *Lactobacillus reuteri* advertised by the formula manufacturer in these samples. When we eliminated this probiotic species from our analysis, there was no qualitative difference in remaining bacterial levels between probiotic-containing and non-probiotic containing samples (Suppl Figure 10b)

## Discussion

More than 78 million families worldwide feed their infants at least partially by bottle^32^making bottle feeding is vital to infant care and equal-opportunity parenting. Surprisingly little is known about its impact on families in terms of financial, emotional and logistical burden, and the evidence underlying recommendations on milk storage remains weak. In our dataset, we found that formula feeding poses the greatest financial burden, while human ilk in bottles is associated with emotional, logistical, and time costs. Given the difficulty of early parenthood, support for parents of infants should integrate an understanding of these challenges. Indeed, our finding that almost half of families discarded milk at least once per day, and that 84% of survey respondents would prefer to discard less milk, underlines the need for more robust data on the necessity of discarding milk for safety reasons.

A body of literature examining the bacteriological composition of fresh (pre-feeding) human milk stored under different conditions has demonstrated bacteriostatic effects of human milk^33–39^, with minimal to no bacterial growth under refrigeration conditions. Thus, storage of fresh human milk at 4°C for up to 4 days has become a standard practice and is compliant with CDC guidelines^6^.

However, to our knowledge, our study represents the largest and most comprehensive examination of bacterial burden in leftover infant milk ***after*** feeding. An undergraduate thesis with 6 infants did address the question of leftover human milk, and reported a range of 762-36,000 CFU/ml at 4°C, which is in the lower range of our findings under similar conditions^9^. In a dissertation from Japan with 11 infant-mother pairs, post-feeding human milk was incubated for 3 and 12h at 4°C, and mean bacterial CFUs before and after feeding were again in the range of our observations, with no relevant difference between pre- and post-feeding milk, and no statistically relevant increase over time occurred^11^. Albeit not peer-reviewed, both publications are in line with our observations and, like our data, support the notion that bacterial growth in leftover infant milk refrigerated up to 8 h is negligible.

In real-world circumstances, keeping a leftover bottle at 4°C may not always be feasible. We therefore explored, for the first time, bacterial loads in leftover milk stored at room temperature. A maximally conservative reading of our data supports the hypothesis that such milk could be safely fed to infants up to 4h post initial feeding, which is the timeframe most likely practically relevant for care givers.

We found no peer reviewed literature examining formula leftover after infant consumption. However, two studies quantified oral bacterial transfer by having adult volunteers suck on artificial bottle nipples^8,11^. In the first, four adults drank infant formula from a bottle through an artificial nipple and the remaining milk was refrigerated for three hours. There was no significant increase in bacteria numbers during storage and the bacterial species composition mirrored the oral flora of the adult participants. In another study, 13 adults repeated this procedure, and no significant bacterial growth was observed upon milk storage at 4°C for 4, 23, and 24 h. The CFU levels were in the lower range of our observations from milk remaining in the bottle after infant feeding. These differences may be due to higher sucking and drinking efficiencies by adults, feeding may influence the transfer of oral flora to the bottled milk. We are not aware of any peer reviewed literature on leftover infant formula kept at room temperature. Although conclusions from our data cannot serve as general recommendations, our findings do support the safety of reoffering leftover infant formula, even when stored up to eight hours post initial feeding at room temperature.

### Strengths and Limitations

Our study is the largest dataset so far on this topic, the first to comprehensively query parents on burdens associated with bottle feeding, the first to systematically compare both formula and human milk storage and the first to longitudinally explore bacterial growth in leftover milk kept at room temperature.

Since our work was conducted in a high-income country, caution should be exercised in transferring these findings to lower resource settings.

Another limitation is that, as participants were aware of the topic of this study, they may have been more rigorous in following hygiene rules than they are in their daily bottle preparation routines.

Importantly, our aim was to gather data on full-term, healthy infants in a non-hospital setting. A much more stringent approach to bottle hygiene and leftover milk disposal must be employed with preterm, immunosuppressed, or hospitalized infants.

While MALDI-TOF mass spectrometry provides reliable bacterial species identification, this method cannot detect viral contamination or certain fastidious pathogens^40^. Furthermore, we chose 20°C as “room temperature” based on typical indoor conditions^41–45^. In certain climates and seasons, bacterial growth may be faster. Additionally, bottles in real-world use may experience temperature fluctuations that were not replicated under controlled laboratory conditions.

## Conclusion

Our findings provide empirical data to inform future evidence-based analyses and refinements of existing guidelines. Under the conditions tested, leftover formula and human milk showed minimal bacterial growth over clinically relevant timeframes, suggesting that current discard recommendations may be more conservative than necessary for healthy, full-term infants in home settings.

## Supporting information

eFigure 1

eFigure 2

eFigure 3

eFigure 4

eFigure 5

eFigure 6

eFigure 7

eFigure 8

eFigure 9

eFigure 10

eTable 1

eTable 2

eTable 3

## Data Availability

All data produced in the present study are available upon reasonable request to the authors

## Acknowledgements

We gratefully acknowledge the infants and families that participated in this study. We are grateful to Dr. Thomas Mueller, Damaris Werner and Romy Cappenberg for their help in recruiting study participants. We also thank Dr. Emily Oster and the entire ParentData community, whose support was invaluable in bringing this work to fruition.

