## Supplementary material for "Leftover Infant Milk After Bottle Feeding: Parental Practices and Microbiological Findings": eFigure 1

**Online-only supplemental Figures**

eFigure 1 Bottle brands and feeding length

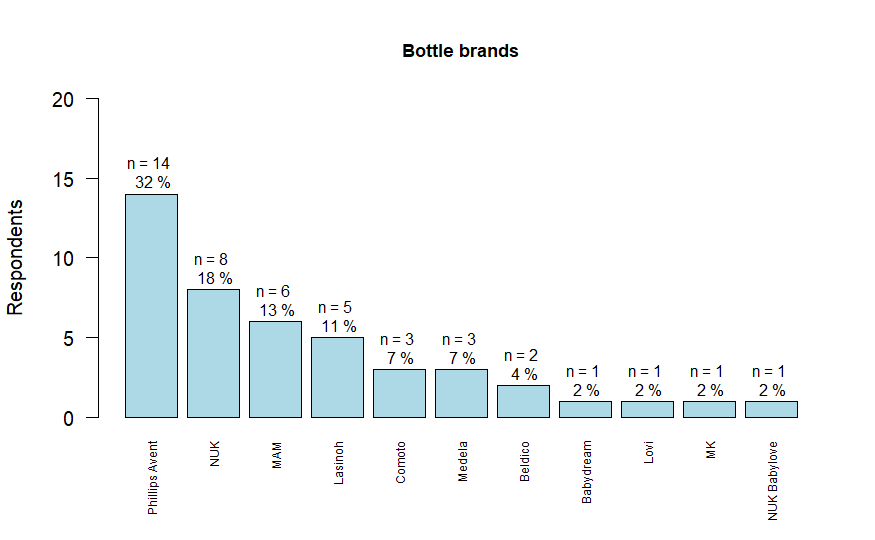

eFigure 1a: Bottle types used by families in microbiology of leftover milk cohort.

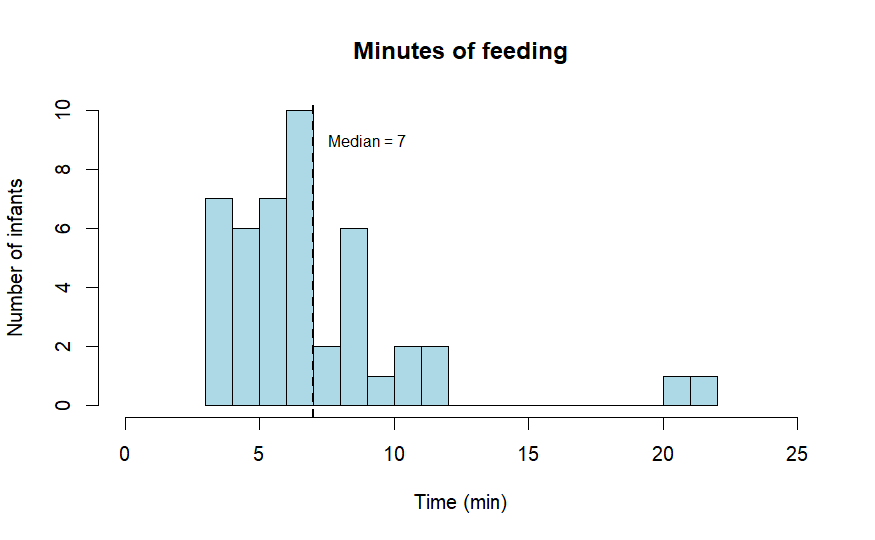
eFigure 1b: Length of feeding from bottle in microbiology of leftover milk cohort (in minutes).
