## Supplementary material for "Leftover Infant Milk After Bottle Feeding: Parental Practices and Microbiological Findings": eFigure 2

**Online-only supplemental Figures**

eFigure 2 Distribution of feeding types among survey respondents

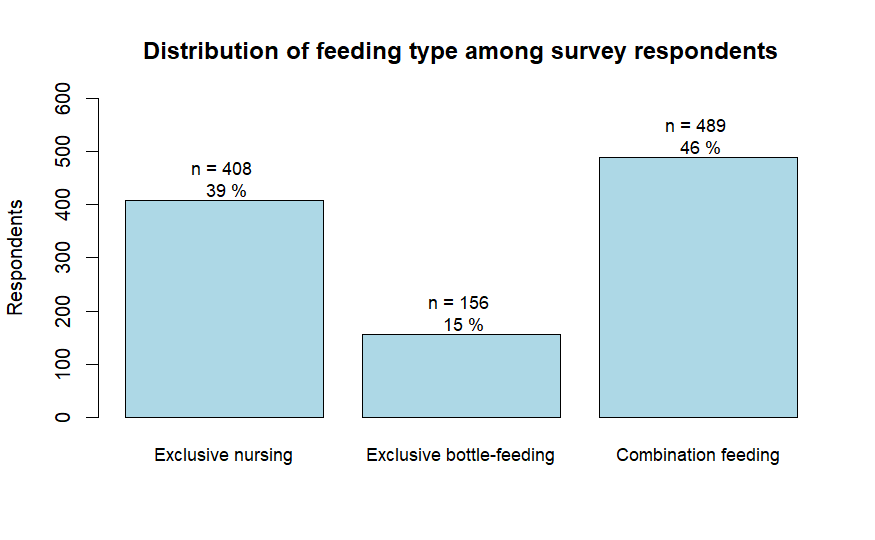

eFigure 2: Distribution of feeding types among survey respondents.
