## Supplementary material for "Leftover Infant Milk After Bottle Feeding: Parental Practices and Microbiological Findings": eFigure 3

**Online-only supplemental Figures**

eFigure 3 Duration before re-offering of milk

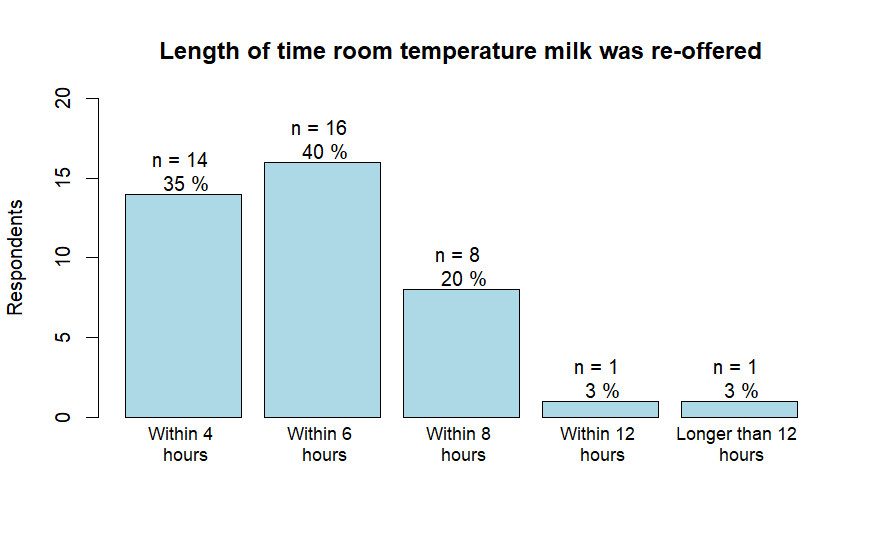

eFigure 3a: Duration milk leftover after the initial feeding was re-offered by parents if kept in the intervening period at room temperature.

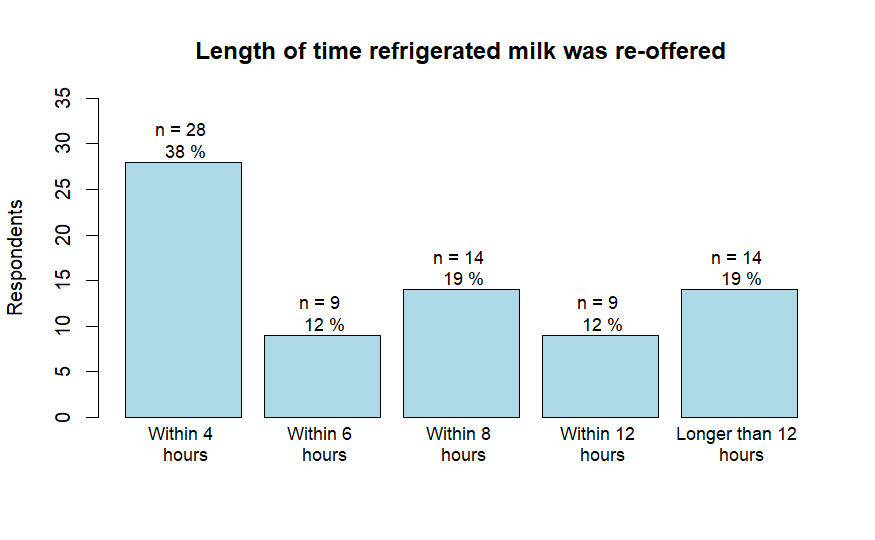

eFigure 3b: Duration milk leftover after the initial feeding was re-offered if kept in the intervening period at room temperature.
