## Supplementary material for "Leftover Infant Milk After Bottle Feeding: Parental Practices and Microbiological Findings": eFigure 4

**Online-only supplemental Figures**

eFigure 4 Types of water used for formula preparation and frequency of bottle sterilization

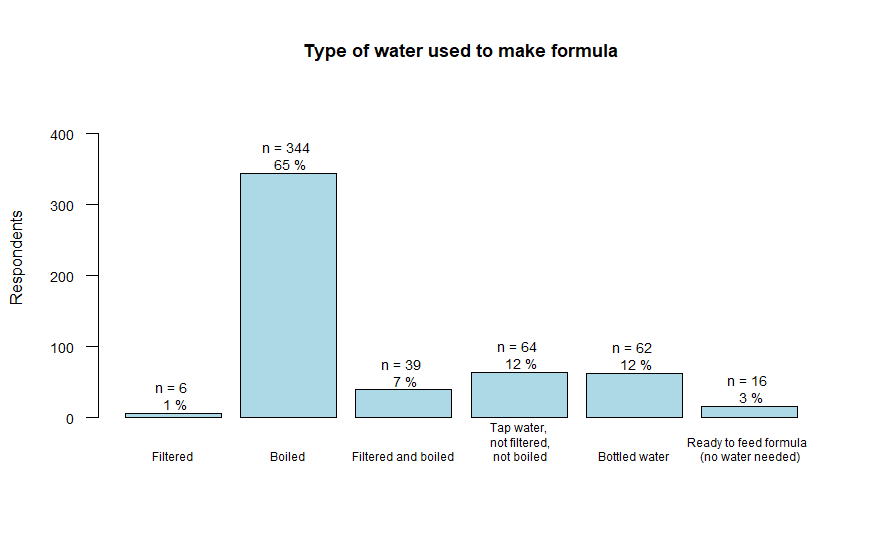

eFigure 4a: Water used to make formula by surveyed parents (n=531).

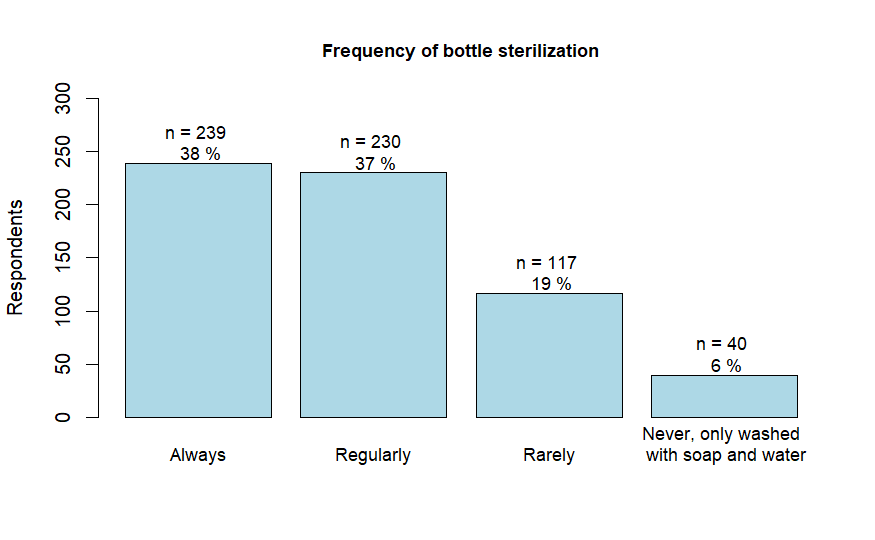

eFigure 4b: Frequency of bottle sterilization reported by the surveyed parents (n=626)
