## Supplementary material for "Leftover Infant Milk After Bottle Feeding: Parental Practices and Microbiological Findings": eFigure 5

**Online-only supplemental Figures**

eFigure 5 Association Between Feeding Duration and Bacterial Levels
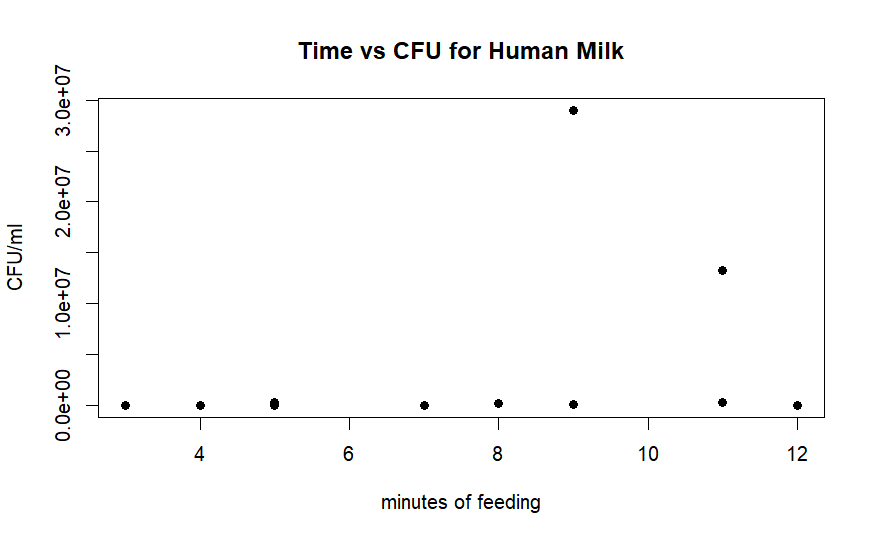

*eFigure 5a: Relationship between time (in minutes) spent drinking pumped human milk from bottle and CFU/ml in leftover milk.*

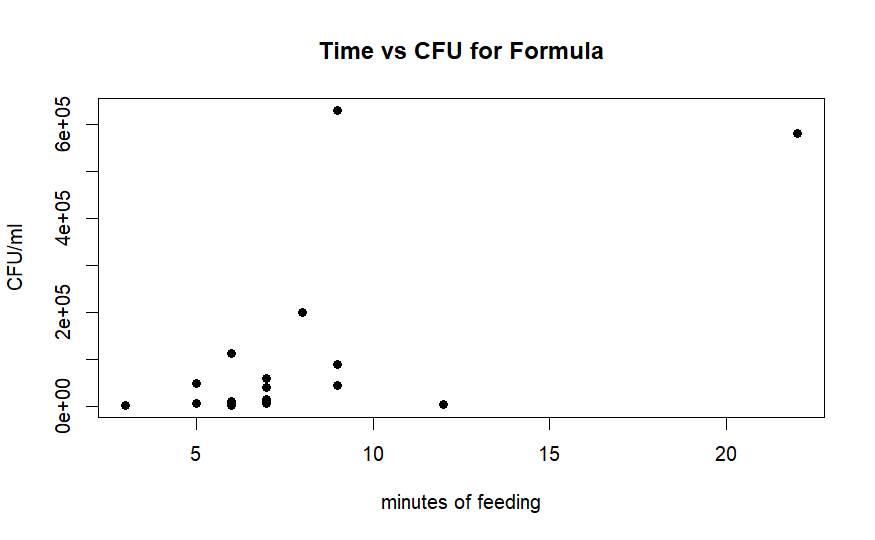

*eFigure 5b: Relationship between time (in minutes) spent drinking formula from bottle and CFU/ml in leftover milk.*
