## Supplementary material for "Leftover Infant Milk After Bottle Feeding: Parental Practices and Microbiological Findings": eFigure 6

**Online-only supplemental Figures**

eFigure 6 Bacterial levels in Leftover Milk by Bottle Material (Glass vs Plastic)

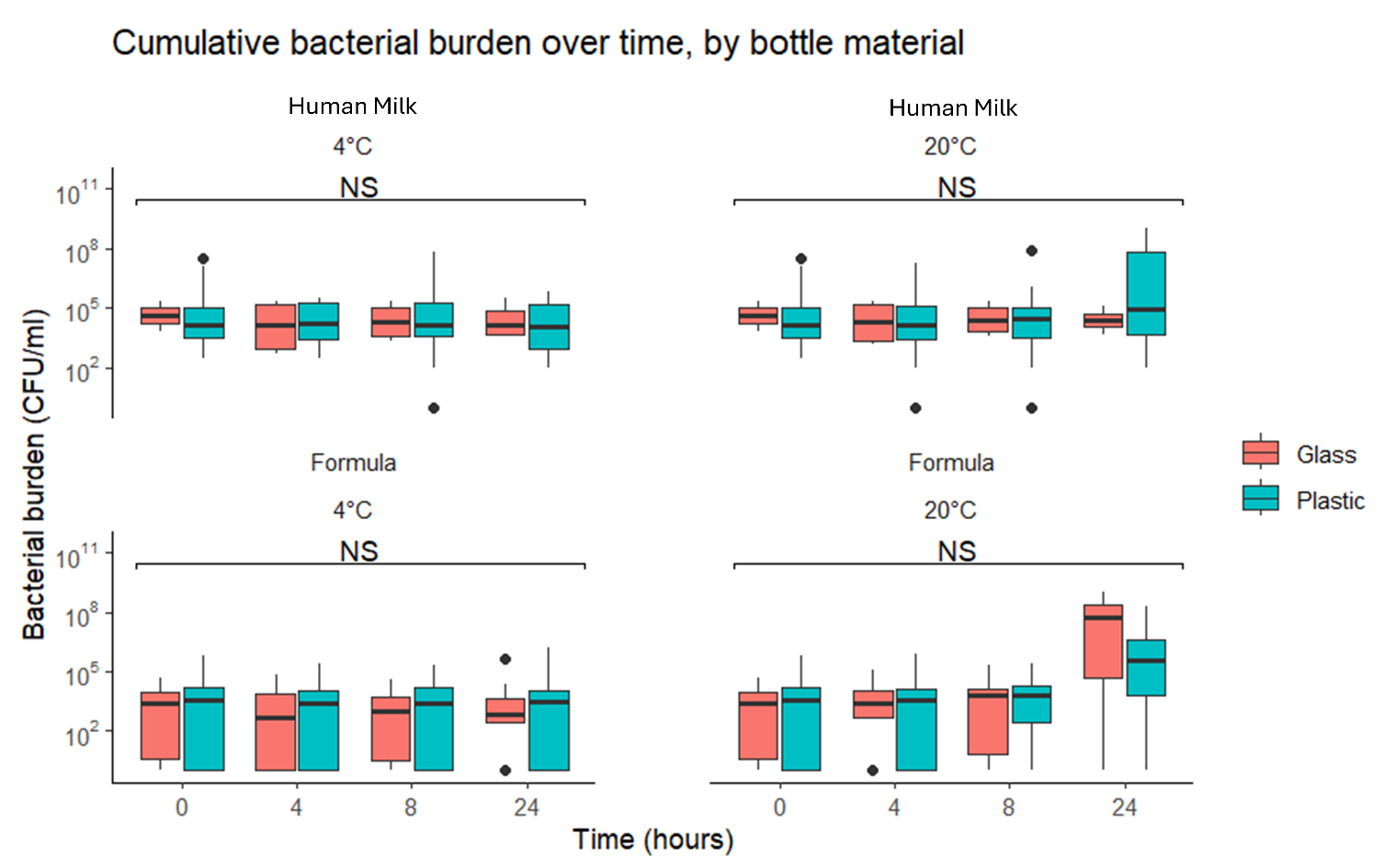

eFigure 6: Relationship between bottle material and CFU/ml. Glass n=9 and plastic n=29. Test used: Pairwise Mood’s median test between all subgroups, with p-values adjusted using the Holm-Bonferroni correction.
