## Supplementary material for "Leftover Infant Milk After Bottle Feeding: Parental Practices and Microbiological Findings": eFigure 7

**Online-only supplemental Figures**

eFigure 7 Bacterial levels in Leftover Milk by Infant Sex and Age

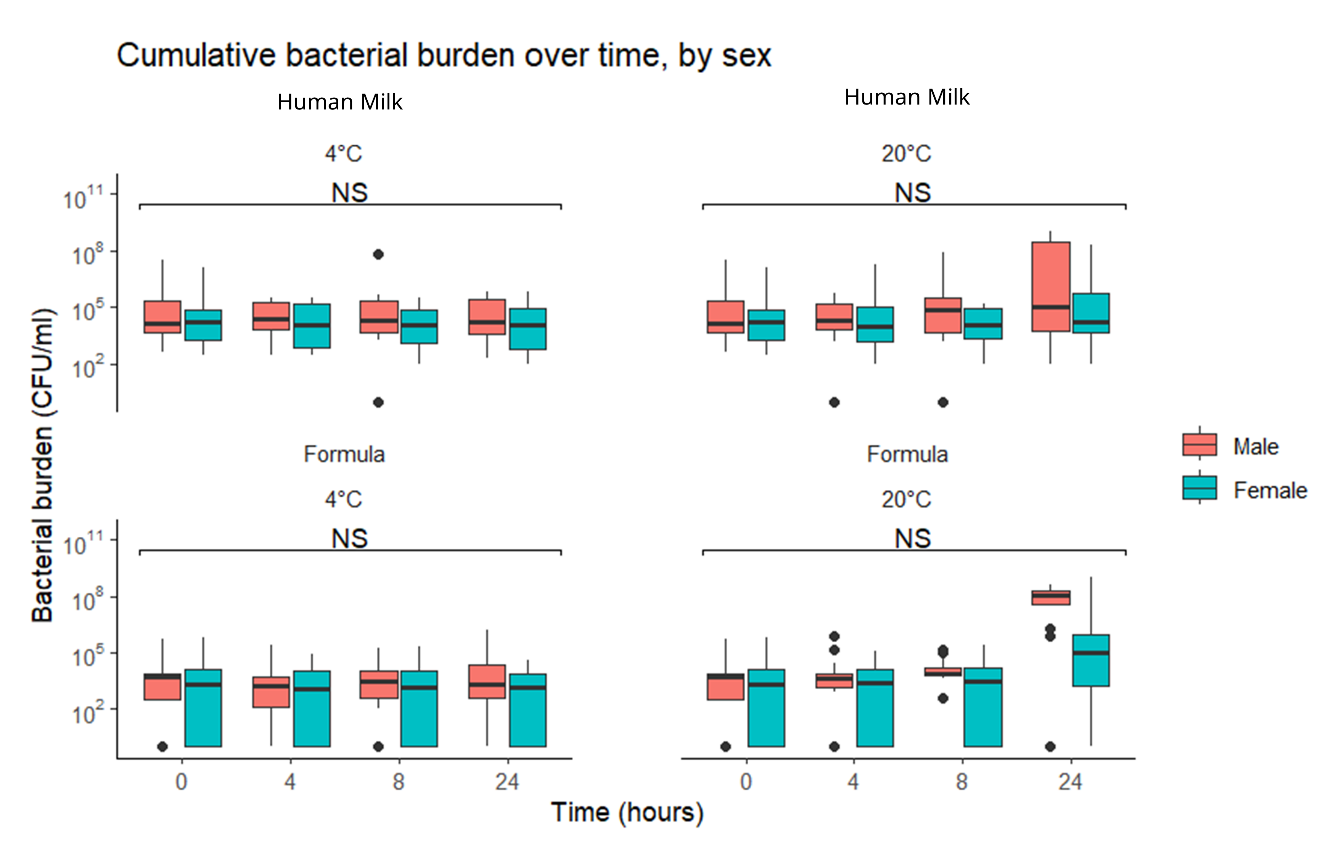

eFigure 7a: Relationship between infant sex and CFU/ml. Male n=21, Female n=23. Test used: Pairwise Mood’s median test between all subgroups, with p-values adjusted using the Holm-Bonferroni correction.

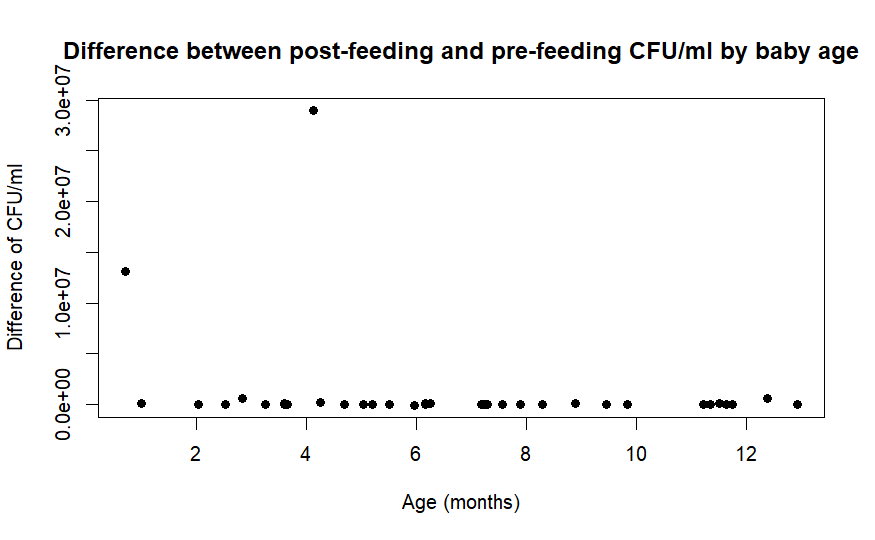

eFigure 7b: Difference in pre-and post-feeding CFU mapped by age of infant. Each dot represents one infant, age is plotted against bacterial CFUs.
