## Supplementary material for "Leftover Infant Milk After Bottle Feeding: Parental Practices and Microbiological Findings": eFigure 8

**Online-only supplemental Figures**

eFigure 8 Post-Feeding Milk Bacterial Species Categorized by Source

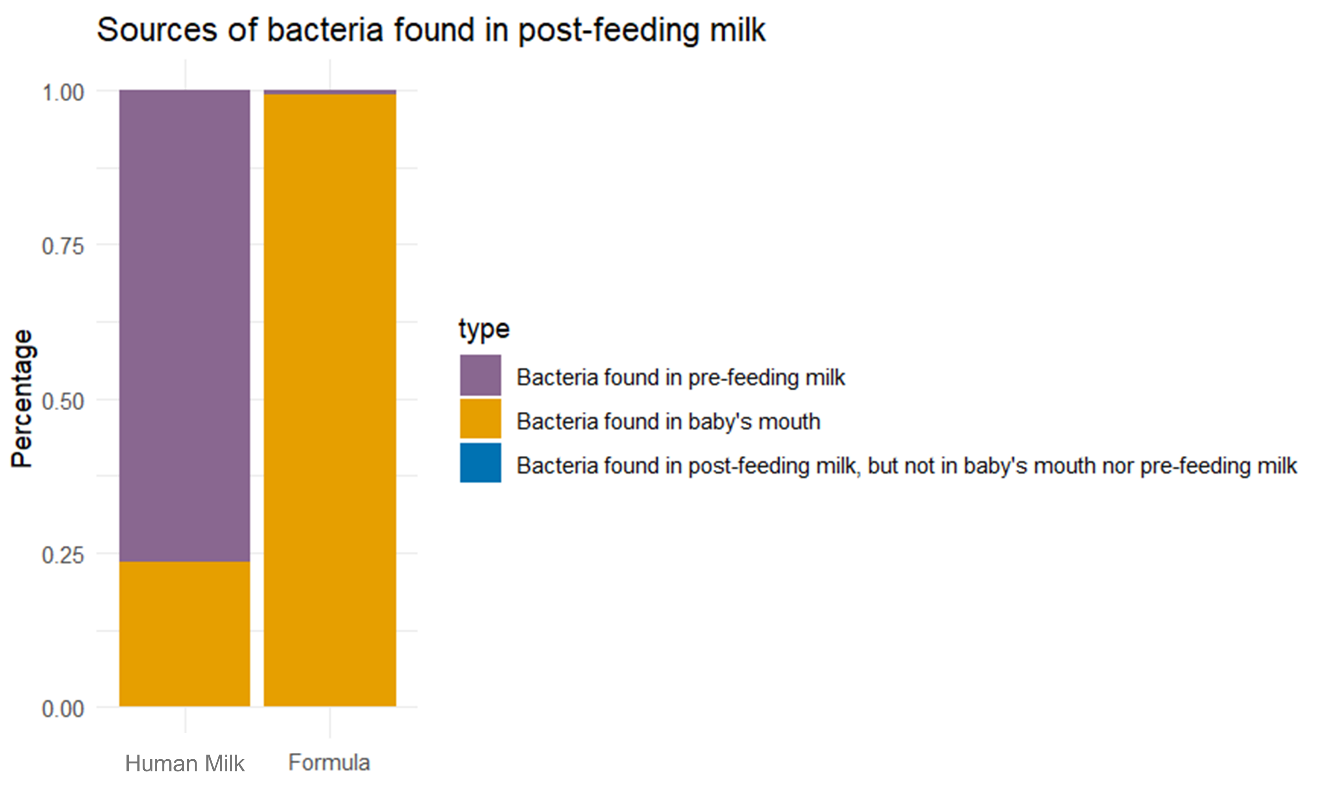

eFigure 8: Bacteria in post-feeding milk, graphed by source. For each bacterial species identified in the bottle post-feeding, source was defined as “baby’s mouth” (yellow) if the species was present in a buccal swab, as “pre-feeding milk” (purple) if the bacterial species was present in the sample removed from bottle before feeding commenced, or as neither (blue).
