## Supplementary material for "Leftover Infant Milk After Bottle Feeding: Parental Practices and Microbiological Findings": eFigure 9

**Online-only supplemental Figures**

eFigure 9 Sources of Bacteria Post-Feeding Milk by Individual Infant

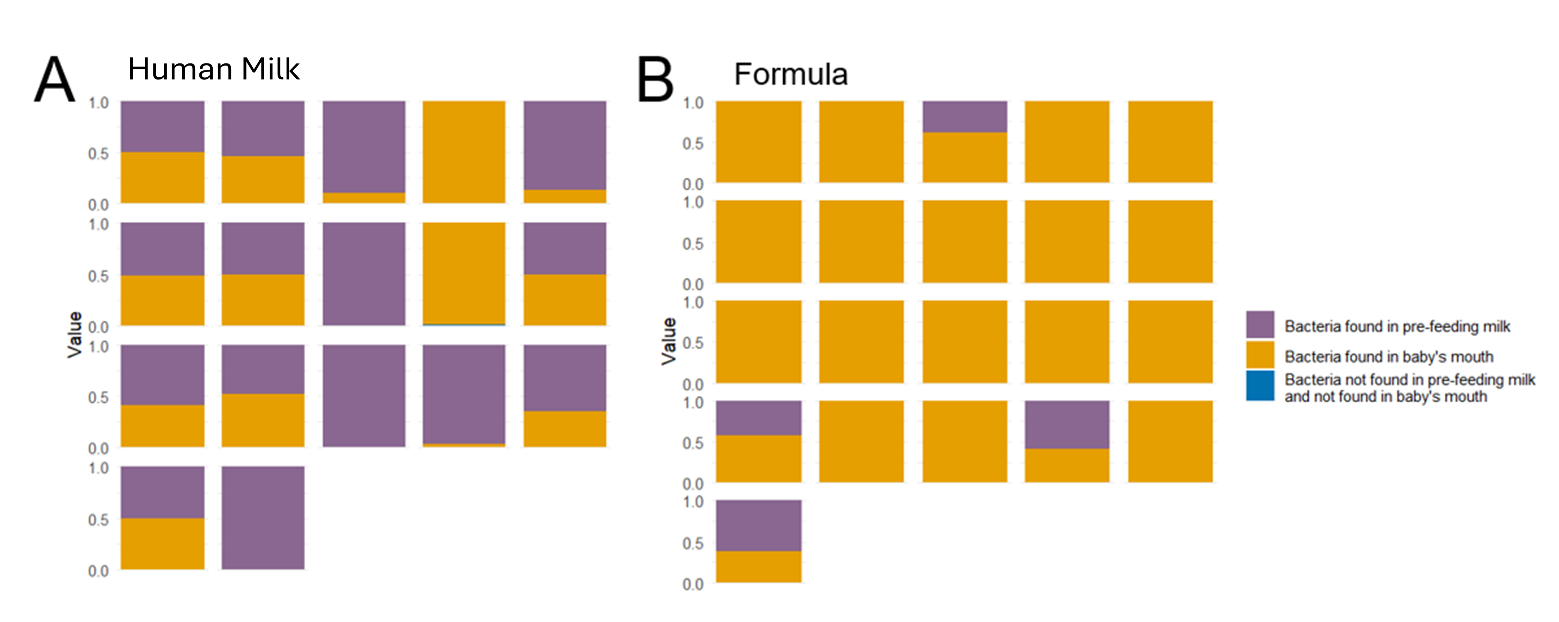

eFigure 9: Source of bacteria found in post-feeding milk, by individual infant. Each box represents the bacterial composition of the post-feeding bottle of one infant, colors reflect the source of the bacterial species identified. Purple= bacterial species found in the pre-feeding bottle, yellow = bacterial species identified in the infant’s buccal swab, blue= bacteria found in neither.
