## Supplementary material for "Leftover Infant Milk After Bottle Feeding: Parental Practices and Microbiological Findings": eFigure 10

**Online-only supplemental Figures**

eFigure 10 Bacterial Burden in Formula Fed to Infants: Unboiled Water and Probiotic Formulas

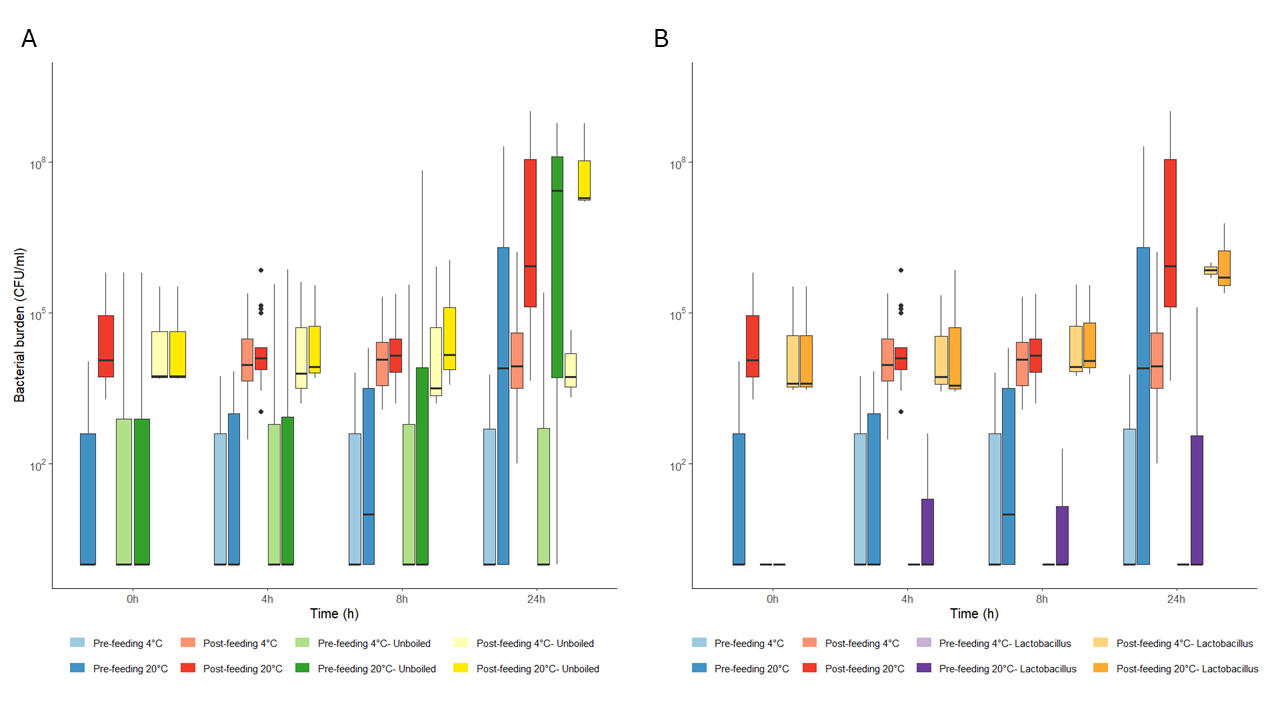

*eFigure 10: (A) Bacterial burden in CFU/ml in the milk of n=3 infants fed with formula made with water that had not being boiled. (B) Bacterial burden in CFU/ml in n=3 infants fed formula containing probiotic cultures. The species of* Lactobacillus *advertised by the manufacturer was removed from the analysis, and only bacterial species non-endogenous to the formula are shown. Bars display median plus interquartile range, n.s.: not significant, CFU: colony forming units, h: hours, °C: degrees celsius,*
