## Supplementary material for "Leftover Infant Milk After Bottle Feeding: Parental Practices and Microbiological Findings": eTable 1

**Online-only supplemental Tables**

eTable 1: List of bacterial species identified in human milk samples

| **Bacteria species** | # of infants with at least one sample containing this bacterial species | Percentage (%) |
| --- | --- | --- |
| ***Staphylococcus epidermidis***  ***Streptococcus-mitis***  ***Rothia mucilaginosa***  ***Staphylococcus aureus***  ***Serratia marcescens***  ***Bacillus cereus***  ***Streptococcus salivarius***  ***Staphylococcus warneri***  ***Enterococcus faecalis***  ***Acinetobacter iwofii***  ***Klebsiella oxytoca***  ***Neisseria subflava***  ***Acinetobacter baumanii***  ***Staphylococcus lugdunesis***  ***Streptococcus sp.***  ***Pseudomonas sp.***  ***Escherichia hermanii***  ***Moraxella oslonensis***  ***Cryseobacterium indologenes*** | 15  11  4  3  3  2  2  2  2  1  1  1  1  1  1  1  1  1  1 | 88  65  24  18  18  12  12  12  12  6  6  6  6  6  6  6  6  6  6 |
