## Supplementary material for "Leftover Infant Milk After Bottle Feeding: Parental Practices and Microbiological Findings": eTable 2

eTable 2: List of bacterial spices identified in formula samples

| **Bacterial species** | # of infants with at least one sample containing this bacterial species | Percentage (%) |
| --- | --- | --- |
| ***Streptococcus mitis***  ***Bacillus cereus***  ***Rothia mucilaginosa***  ***Neisseria subflava***  ***Streptococcus salivarius***  ***Neisseria sp.***  ***Staphylococcus epidermidis***  ***Staphylococcus hominis***  ***Bacillus firmus***  ***Staphylococcus aureus***  ***Brevibacillus parabrevis***  ***Non-fermenter***  ***Coagulase negative staphylococcus***  ***Acinetobacter pittii***  ***Neisseria perflava***  ***Neisseria mucosa***  ***Chrysobacterium sp.***  ***Micrococcus luteus***  ***Gemella sp.***  ***Acinetobacter junii***  ***Bacillus megaterium***  ***Bacillus sp.***  ***Bacillus horneckiae*** | 21  8  7  5  5  3  3  3  2  1  1  1  1  1  1  1  1  1  1  1  1  1  1 | 100  38  33  24  24  14  14  14  10  5  5  5  5  5  5  5  5  5  5  5  5  5  5 |
