## Supplementary material for "Leftover Infant Milk After Bottle Feeding: Parental Practices and Microbiological Findings": eTable 3

**Online-only supplemental tables**

eTable 3: List of bacterial species identified in infant buccal swabs

| **Bacterial species** | # of infants with at least one sample containing this bacterial species | Percentage (%) |
| --- | --- | --- |
| ***Streptococcus-mitis Group***  ***Rothia mucilaginosa***  ***Neisseria subflava***  ***Streptococcus salivarius***  ***Neisseria sp.***  ***Staphylococcus epidermidis***  ***Staphylococcus hominis***  ***Staphylococcus aureus***  ***Gemella sp.***  ***Coagulase negative staphylococcus***  ***C. albicans***  ***Citrobacter sp.***  ***Gemella haemolysans***  ***Haemophilus influenzae***  ***Haemophilus parainfluenzae***  ***Haemophilus spp.***  ***Klebsiella oxytoca***  ***Klebsiella pneumonia***  ***Micrococcus luteus***  ***Neisseria mucosa***  ***Neisseria perflava***  ***Serratia marcescens***  ***Staphylococcus haemolyticus***  ***Streptococcus parasanguinis***  ***Streptococcus sp.***  ***Streptococcus vestibularis*** | 35  16  9  9  5  5  4  3  2  2  1  1  1  1  1  1  1  1  1  1  1  1  1  1  1  1 | 92  42  24  24  13  13  11  8  5  5  3  3  3  3  3  3  3  3  3  3  3  3  3  3  3  3 |
